# Systemic and Genetic Risk Factors for Reticular Macular Disease and Soft Drusen in Age-Related Macular Degeneration

**DOI:** 10.1101/2021.09.27.21263712

**Authors:** Robert J. Thomson, Joshua Chazaro, Oscar Otero-Marquez, Gerardo Ledesma-Gil, Yuehong Tong, Arielle C. Coughlin, Zachary R. Teibel, Sharmina Alauddin, Katy Tai, Harriet Lloyd, Maria Scolaro, Arun Govindaiah, Alauddin Bhuiyan, Mandip S. Dhamoon, Avnish Deobhakta, Jagat Narula, Richard B. Rosen, Lawrence A. Yannuzzi, K. Bailey Freund, R. Theodore Smith

## Abstract

**Purpose:** Soft drusen and subretinal drusenoid deposits (SDD) aka reticular macular disease (RMD) characterize two pathways to advanced age-related macular degeneration (AMD). We propose these pathways are distinct diseases, with distinct genetic risks, serum risks and associated systemic diseases.

**Methods:** 126 Subjects with AMD had: retinal imaging for RMD status, serum risks, genetic testing, and histories of cardiovascular disease (CVD) and stroke.

**Results:** 62 subjects had RMD, 64 were nonRMD (drusen only), 51 had CVD or Stroke. RMD correlated significantly with: *ARMS2* risk allele (p= 0.019); lower mean serum HDL (61±18 vs. 69±22 mg/dl, p= 0.038, t test); CVD and troke (34/51 RMD, p= 0.001).NonRMD correlated/trended with *APOE2* (p= 0.032) and *CETP* (p= 0.072) risk alleles. 97 subjects total had some drusen, which correlated with *CFH* risk (p= 0.016). Multivariate independent risks for RMD were: CVD and Stroke (p= 0.008), and *ARMS2* homozygous risk (p= 0.038).

**Conclusion:** The RMD and soft drusen AMD pathways have distinct systemic associations, serum and genetic risks. RMD is associated with CVD and stroke, *ARMS2* risk, and lower HDL; drusen with CFH risk and two lipid risk genes. These pathways appear to be distinct diseases leading to advanced AMD.

**Summary Statement:** Two phenotypes of age-related macular degeneration, soft drusen and reticular macular disease (the combination of subretinal drusenoid deposits and choriocapillaris insufficiency), are shown here to have distinct systemic vascular, serum, and genetic risks. These findings support the concept that these phenotypes actually represent distinct disease processes.

---

Multiple systemic, environmental and genetic risk factors have been identified for the complex disease of Age-related macular degeneration (AMD). However, assessment of the disease impact of variables from so many sectors is difficult, and the pathogenic mechanisms driving the disease itself are largely uncertain. We herein suggest that this problem be broken into two largely independent disease pathways, which could be more easily analyzed separately. These two pathways involve distinct lipid-laden deposits; one with lesions of soft drusen beneath the retinal pigmented epithelium (RPE), and one with subretinal drusenoid deposits (SDD) above the RPE^1^ (**Figure 1)**. Both deposits can occur in the same eye (**Figure 1B)**, and both can progress to advanced AMD,^2-4^ of which SDD, aka reticular pseudodrusen (RPD), confers twice the risk.^5,6,7^

**Figure 1.**
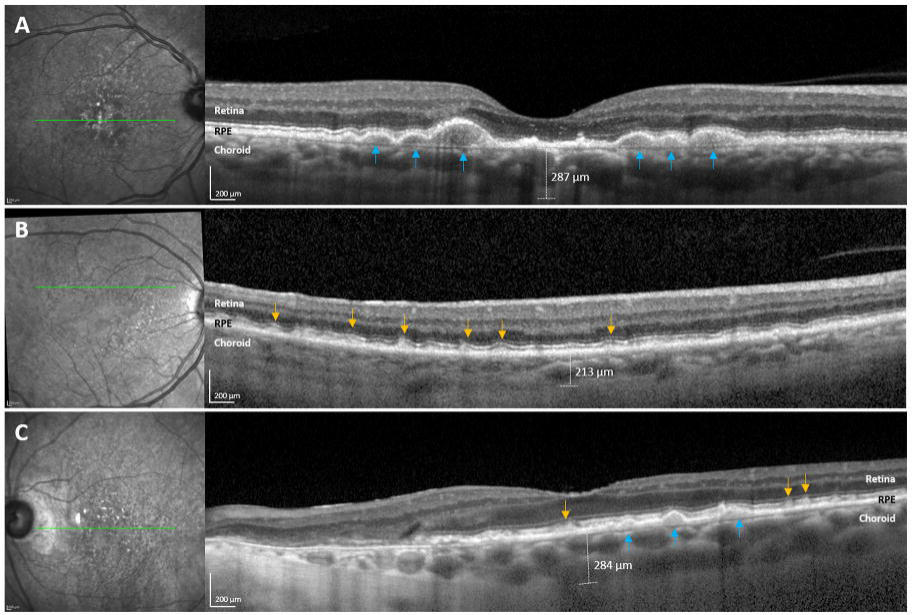
Multimodal Imaging of Soft Drusen and Reticular Macular Disease (RMD) in Age-related Macular Degeneration (AMD). **Left**. Near infrared reflectance (NIR) images. **Right**. Spectral domain optical coherence tomography (SD-OCT) images. The green lines on the NIR images denote the location of the corresponding B-scans on SD-OCT. **A. Soft drusen. NIR** lesions of soft drusen have a hyperreflectant appearance. **SD-OCT**. Soft drusen (blue arrowheads) lie between the basal lamina of the retinal pigment epithelium (RPE) and the inner collagenous layer of Bruch’s membrane (BrM), which is seen intermittently where the RPE is elevated. The choroidal thickness in this case is 287 microns. **B. Mixed drusen and RMD. NIR** lesions of RMD are fairly homogeneous and moderately hypo-reflectant in a compact distribution with well-defined borders that is pathognomonic for RMD. **SD-OCT**. SDD Stage II lesions between the RPE and ellipsoid zone (EZ) are large enough to distort the EZ (yellow arrowheads). The choroidal thickness in this case is 284 microns. **C. Pure RMD. NIR** shows pathognomonic hypo-reflectant lesions, more numerous superiorly as is usually the case. **SD-OCT**. A few stage I lesions not distorting the overlying EZ are seen between the stage II SDD. In this pure RMD (no soft drusen) the choroidal thickness is 213 microns, thinner than that in A and B, the soft drusen and mixed phenotypes.

An important further consideration is functional: the RPE is supplied by the choroidal vasculature/choriocapillaris (CC).^8^ CC insufficiency uniformly accompanies SDD on indocyanine green angiography^9^ and non-invasive optical coherence tomography angiography (OCTA) ^10^ often accompanied by choroidal thinning and loss of choroidal volume.^11^ SDD are topographically related to choroidal watershed zones, suggesting hypoxia in their pathogenesis.^12^ Hence CC insufficiency and SDDs have logically been combined together as the reticular macular disease (RMD) phenotype of AMD^9^, and we will refer to this phenotype unambiguously as SDD or RMD.

We propose these hypotheses to be explored initially herein and as a working basis for productive future research in AMD.

**Hypotheses: 1**. The SDD and soft drusen pathways do not have the same systemic associations, serum risks or genetic risks. **2**. These pathways are in fact distinct diseases that only merge in their common outcome in what is truly a single disease, advanced AMD (atrophic and/or exudative). **3**. Patient care could be personalized by consideration of the risk factors pertinent to the disease pathway operative in a given individual. Research on the pathogenesis of AMD could be well-served by identifying and differentiating those risk factors that apply specifically to only one or the other pathway, enabling focused attention to specific risks and single pathways, with corresponding simplification and higher chance of success.

**Hypothesis 1** will be explored, amplified and supported in this paper.

**Hypothesis 2**. Evidence is presented here in the categories mentioned that SDD/RMD and soft drusen are hallmarks of distinct diseases. Broad and persuasive support, to be summarized, is already established.

**Hypothesis 3** is theoretically self-evident, given **Hypotheses 1 and 2**,

Regarding **Hypothesis 2:** established multimodal imaging and anatomic distinctions between the phenotypes include: scanning laser ophthalmoscopy (SLO), with autofluorescence (AF) and near-infrared reflectance (NIR) imaging, which clearly identifies SDD as hypoautofluorescent on AF and hyporeflectant on NIR,^9^ properties not shared by drusen; histopathological identification of SDD^13^ in the subretinal space, with drusen in the sub-RPE space, confirmed *in vivo* by spectral domain OCT (SD-OCT),^14^ with choroidal thinning commonly found with SDD; and topographic localization of SDD with rod photoreceptors.^15^ SDD are associated epidemiologically with female gender and decreased longevity,^5^ systemically with coronary artery disease (CAD) in small case-control studies,^2 3^ since confirmed with OCTA documentation of CC insufficiency,^16^ and with cardiovascular cause of death in donors.^17^ SDD carry a worse prognosis for progression to advanced AMD^6^ and differing associated subtypes of advanced AMD (both exudative, type 3 choroidal neovascuarization^18^ and non-exudative, multilobular geographic atrophy^7^), biochemical composition,^1^ and functional impact on rod-mediated dark adaption.^19^

Herein, we extended these disease distinctions with data on AMD risk alleles, serum risk markers, and associations with vascular disease, as well as other demographic, ocular and systemic features. We classified AMD patients into RMD (RMD present, either eye, +/-drusen) with obligate underlying CC deficiency or non-RMD (large soft drusen only) by retinal imaging for comparison

## Methods

This was a multicenter prospective study conducted at two tertiary vitreoretinal referral centers in New York City, USA: Vitreous Retina Macula Consultants of New York (LY, KBF), and Department of Ophthalmology, New York Eye and Ear Infirmary of Mount Sinai School of Medicine (MSSM) (RBR, RTS), from January 2019 to January 2021. The institutional review board of MSSM approved the study, which adhered to the tenets of the Declaration of Helsinki. ClinicalTrials.gov Identifier: NCT04087356.

### Inclusion and Exclusion Criteria

All patients older than 50 years, diagnosed with RMD/SDD and/or large soft drusen lesions of AMD in either eye, who signed informed consent, and completed a study-related questionnaire were included. Exclusion criteria were patients with other retinal degeneration and vascular diseases, prior retinal surgery (except intravitreal injections), poor-quality imaging and/or inconclusive medical or macular diagnoses.

### Patient Evaluation

Work-up included best-corrected visual acuity, slit lamp biomicroscopy, intraocular pressure (mmHg), blood samples for 9 AMD risk alleles and vascular disease risk factors, age, gender, race/ethnicity, relevant family history, smoking, medication, and medical/ocular history specifically covering stroke and these heart disease histories: myocardial infarction, coronary artery bypass grafting (CABG), angina, arrhythmia, positive stress test, positive cardiac catheterization, stent, valve disease and congestive heart failure (CHF) of any cause. Cases confirmed against medical records were included in the cardiovascular (CVD) and stroke group.

### Imaging

Volume SD-OCT scans (27 lines, automated retinal tracking, 16 scans averaged per line), and *en face* AF and NIR scans (both 30 degrees) centered on the fovea on the Heidelberg Spectralis HRA + OCT (Heidelberg engineering, Heidelberg, DE).

### Image Analysis

SDD were identified on SD-OCT imaging as lesions of intermediate reflectivity above the RPE followed a published protocol^2^ for presence and most advanced stage independently and then to consensus by two retina specialists (GLG, OOM). Any differences were resolved by a senior grader (RTS). AF and NIR images were used to confirm the presence of SDD as fairly uniform hypoautofluorescent or hyporeflective lesions in compact distributions with well-defined borders.^9^ RMD status was determined by the presence of SDD alone, relying on significant literature uniformly demonstrating CC insufficiency ^9,10 12,20^ Soft drusen were identified on SD-OCT imaging as lesions of intermediate reflectivity on Bruch’s membrane (BrM) elevating the RPE (**Figure 1**). Pigment epithelial detachments (PEDs) were included with soft drusen. Other drusen phenotypes of lesser prevalence (calcified drusen, cuticular drusen) were not included. A large soft drusen was required to be >125 microns in diameter. Choroidal thickness (CTh) was measured on a central SD-OCT scan (**Figure 1**).

### Serum CVD Risk Markers Analysis

Blood samples for risk biomarkers of CVD^21^ (high-density lipoprotein cholesterol [HDL-c], low-density lipoprotein cholesterol [LDL-c], triglycerides [TG], total cholesterol [TC] and high sensitivity C-reactive protein [hsCRP]) were rapidly centrifuged at 1800 g for 10 minutes and refrigerated. Lipid levels were measured (Quest Diagnostics, Teterboro, NJ) by spectrophotometry, and plasma levels of hsCRP, by Immunoturbidimetric Assay (Orion Diagnostica, Finland).

### Genetic Analysis

Blood samples for DNA were immediately stored at - 70°C until shipped on dry ice to LGC Genomics (LGC, Biosearch Technologies, Hoddesdon, UK). Genomic DNA was extracted from peripheral blood leukocytes according to established protocols. Genotyping was carried out using KASP (Kompetitive Allele-Specific PCR, LGC Genomics) genotyping assays designed to each target variant, within 1536 well PCR plates. Fluorescence was detected using the BMG Pherastar (BMG LABTECH GmbH, Ortenberg, DE) and genotyping calls assigned based on cluster plots within the LGC Genomics Kraken software.

The samples were assessed for single-nucleotide polymorphisms in 9 genes associated with AMD: *CFH* (Y402H allele), *ARMS2-HTRA1* (69S allele), *C3, C2/BF* (*BF* rs4151667, which identifies the H10 haplotype of *C2/BF*), cholesteryl ester transfer protein (*CETP)*, hepatic lipase (*LIPC*), tissue inhibitor of metalloproteinase 3 (*TIMP3*), apolipoprotein genes (*APOE2* and *APOE4*). All are risk alleles except for the protective *C2/BF, LIPC* and *APOE4* alleles.^22^

### Statistical Analysis

Experts (AB, AG) used ‘IBM SPSS Statistics version 27’, ‘Waikato Environment for Knowledge Analysis (WEKA) Version 3.8.5’, a data modeling tool, and Microsoft Excel 365. Univariate statistics were chi square for categorical variables, two-tailed t-test for continuous variables, with medians, ranges and/or interquartile ranges (IQRs) for non-normally distributed data. Multivariate regression determined each variable’s significance (p< 0.05) after controlling for all other covariates.

## Results

### Demographics and clinical characteristics

62 subjects had SDD/RMD present (29 pure, without large soft drusen, and 33 mixed cases) and 64, SDD absent (large soft drusen only). Five disagreements were resolved by consensus without arbitration. For the RMD group, 67.7% were females and 32.3% were males. The associations of demographic, ocular and clinical characteristics of patients with or without RMD were not significantly different, except that acetylsalicylic acid (ASA) use was significantly associated with RMD (53/62 RMD vs 30/64 nonRMD, p= 0.007), and mean CTh was less in left eyes with RMD than in nonRMD left eyes (Mean 146±58 vs. 177±59 microns, p**=** 0.003; median 161.5, range 78-228 microns vs. median 177.5, range 90-286 microns) as shown in **Table 1**.

### Serum Risk Markers, CVD and Stroke

Cholesterol panel and hsCRP mean values for RMD and non-RMD subjects were not significantly different, except mean HDL was lower in RMD (mean 61±18; median 57, IQR 19) than non-RMD subjects (mean 69±22, p= 0.038; median 69, IQR 29) as depicted in **Table 2**.

Of the 126 subjects, 51 had CVD or stroke, 34/51 RMD (p= 0.001, chi square).

### Genetics

The *ARMS2* risk allele frequency was significantly higher, 41.6%, in the RMD group than in the nonRMD group (28.5%, OR 1.79, 95% CI 1.15 - 3.03, p= 0.03).

The *CFH* risk allele frequencies were similar in the two groups (56% and 53.2%; p= 0.66).

Dividing the subjects analogously into Drusen (Drusen present, +/-RMD) or nonDrusen (RMD only, no large soft drusen), the *CFH* risk allele frequency was significantly higher, 61.4%, in the Drusen group than in the nonDrusen group (36.8%, OR 2.37, 95% CI 1.16-4.76, p= 0.016). All other findings with this subdivision were NS.

*APOE2* and *CETP* risk allele frequencies were higher in the nonRMD group than in the RMD group (14.3% vs 5.9%, and 43.6% vs 32.5%, p= 0.03 and 0.07, respectively). Allele frequencies and significance of correlations with RMD or nonRMD of haplotype-tagging single-nucleotide polymorphisms (SNPs) from the AMD-associated Loci are shown in **Table 3**.

### Multivariate regression

Multivariate regression found CVD or stroke and *ARMS2* homozygous risk significant for RMD risk (p= 0.008 and p= 0.038, respectively), after controlling for all other parameters. ASA use, a *treatment* for many vascular diseases, was not analyzed for multivariate RMD risk, considering the evident strong association of vascular diseases with RMD/SDD.

## Discussion

The central hypothesis presented in this paper is that the SDD/RMD and soft drusen pathways of AMD are actually two separate, but often co-existing, diseases that may result in a single disease outcome, advanced AMD. The phenomenon of multiple, separate disease pathways resulting in the same poor outcome and organ dysfunction is often the norm in medicine rather than the exception, e.g., the many unique paths to heart failure. As mentioned before, evidence for this pathophysiologic distinction has already accumulated from diverse disciplines: anatomy, histology, imaging, retinal function, clinical outcomes, biochemistry, and choroidal vascular physiology. Indeed, if all these facts had been known at the outset with the discovery of the new lesion of SDD, the recognition of a completely distinct disease process would have likely been clear and immediate.

We have shown herein that the SDD/RMD and soft drusen pathways also differ significantly in their systemic associations, serum risks and genetic risks. In total, we suggest the evidence is persuasive. As a corollary, this construct further supports revising the term AMD to be reserved for the outcomes now known as advanced AMD, with age-related maculopathy (ARMD) applied to any precursor state: RMD, drusen, other.

The *ARMS2* risk allele significantly correlated with reticular macular disease (RMD), the combined phenotype of SDD and choriocapillaris insufficiency, compared to soft drusen; and the *CFH* risk allele significantly correlated with soft drusen compared to RMD. These results replicate our previous discovery that *ARMS2* risk is significantly more correlated with RMD than nonRMD/pure drusen in AMD.^23^ The finding that CFH risk is significantly higher in all subjects with drusen than pure RMD, is new, but is consistent with our 2011 paper. This still leaves the possibility that both genes contribute to both pathways, but CFH weakly to SDD and ARMS2 weakly to soft drusen.

CVD and stroke are the leading causes of death in the developed world. AMD is the leading cause of blindness.^4^ Previous studies of AMD as a whole have failed to find consistent relations between these diseaes^24-26^ and in a large metanalysis, no associations. ^27^ However, SDD and choroidal thinning specifically have been found to be associated with CAD in small studies.^2,3^ SDD also confer decreased longevity,^5^ with vascular death suspected.^2,3^ Histopathology of 1777 donor eyes^17^ found SDD, alone among AMD lesions, were associated with cardiovascular death. We have shown herein additional convincing evidence that CVD and stroke are strongly associated with the SDD/RMD type, not soft drusen.

The lipid dynamics of drusen with esterified cholesterol and SDD with non-esterified cholesterol ^1^ are complex. Lipid in drusen and in the plaques of atherosclerotic disease (ASD) led to a long search for common mechanisms in these lesions. However, it is increasingly clear that drusen accumulation is associated with local blockage by abnormal BrM of normal absorption by the choroidal circulation of lipids secreted by the RPE.^1^ Further, as just reviewed, systemic vascular disease is associated with RMD, not drusen.

The risk marker of low serum HDL for ASD actually runs contrary for AMD, with higher HDL increasing risk.^28^ This apparent paradox may now be explained: prior studies considered only the drusen phenotype of AMD in color photos, and indeed, showed that high HDL is a risk for drusen in AMD.^28^ Low HDL is a risk for CVD and stroke, which we now have found is a risk for RMD, the other AMD pathway. HDL cuts both ways, depending on the pathway.

The many (29/62) cases of pure RMD (without drusen) further suggest that RMD is a distinct retinal pathology. Damage from RMD then can lead, along with that of soft drusen, to advanced AMD, with similar presentations. It should be noted, however, that the lesions of geographic atrophy consequent to drusen or RMD still have distinguishing morphologic^7^ and autofluorescence characteristics,^29^ consistent with different diseases.

The study has several weaknesses. The sample size is relatively small, so the results, from a mostly Caucasian elderly population, require further examination in larger and diverse cohorts e.g., in Asians, with the prevalent polypoidal choroidal vasculopathy form of AMD.^30^ RMD status was determined by the presence of SDD alone, relying for CC insufficiency on the quite consistent literature ^9 12,20^ without direct verification by OCTA; this can be added in future studies. Important drusen phenotypes other than soft drusen, such as calcified drusen and cuticular drusen, have their own identified risk factors, but were not included. Larger studies with greater power will be doubtless be needed to determine the associations of such phenotypes with lesser prevalence. Vascular histories were patient reported and were verified from medical records only in positive cases with stated heart disease or stroke. More detailed data on cardiac and carotid status should be included in future studies to help interpret these results.

Strengths of this prospective study at two tertiary retina referral centers include rigorous patient selection and AMD phenotyping with high quality multimodal imaging for drusen and RMD. It is significant that the associations from 3 new fields, systemic vascular disease, serum risks and genetic risks, further differentiate the drusen and RMD pathways with new data that extend the already broad platform of existing evidence.

## Conclusion

The SDD/RMD and soft drusen pathways, now considered to be components of one disease, AMD, actually appear to be two separate, often co-existing diseases that only become part of a single disease in their common outcome, advanced AMD.

There may be very real practical benefit from recognizing this pathophysiology. As shown here, many AMD patients demonstrate only one of these index lesions. Patient care could be personalized by consideration of the risk factors pertinent to such individuals. Research on the pathogenesis of AMD could also be well-served by identifying and differentiating those risk factors that apply specifically to only one or the other pathway. These distinctions could enable focused attention on specific risks and single pathways, with correspondingly higher chances of success. The specific association between systemic vascular disease and RMD in particular merits further study in the context of the choroidal vascular insufficiency that accompanies SDD and defines RMD.

## Data Availability

n/a

## Acknowledgment

we gratefully acknowledge several helpful conversations with Christine Curcio.

## REFERENCES

1. Oak AS, Messinger JD, Curcio CA. Subretinal drusenoid deposits: further characterization by lipid histochemistry. Retina. 2014;34(4):825–826.

2. Cymerman RM, Skolnick AH, Cole WJ, Nabati C, Curcio CA, Smith RT. Coronary Artery Disease and Reticular Macular Disease, a Subphenotype of Early Age-Related Macular Degeneration. Curr Eye Res. 2016;41(11):1482–1488.

3. Ahmad M, Kaszubski PA, Cobbs L, Reynolds H, Smith RT. Choroidal thickness in patients with coronary artery disease. PLoS One. 2017;12(6):e0175691.

4. Miller JW. Age-related macular degeneration revisited--piecing the puzzle: the LXIX Edward Jackson memorial lecture. Am J Ophthalmol. 2013;155(1):1–35 e13.

5. Klein R, Meuer SM, Knudtson MD, Iyengar SK, Klein BE. The epidemiology of retinal reticular drusen. Am J Ophthalmol. 2008;145(2):317–326.

6. Pumariega NM, Smith RT, Sohrab MA, LeTien V, Souied EH. A prospective study of reticular macular disease. Ophthalmology. 2011;118(8):1619–1625.

7. Xu L, Blonska AM, Pumariega NM, et al. Reticular macular disease is associated with multilobular geographic atrophy in age-related macular degeneration. Retina. 2013;33(9):1850–1862.

8. Hayreh SS. In vivo choroidal circulation and its watershed zones. Eye (Lond). 1990;4(Pt 2):273–289.

9. Smith RT, Sohrab MA, Busuioc M, Barile G. Reticular macular disease. Am J Ophthalmol. 2009;148(5):733–743.

10. Nesper PL, Soetikno BT, Fawzi AA. Choriocapillaris Nonperfusion is Associated With Poor Visual Acuity in Eyes With Reticular Pseudodrusen. Am J Ophthalmol. 2017;174:42–55.

11. Cheng H, Kaszubski PA, Hao H, et al. The Relationship Between Reticular Macular Disease and Choroidal Thickness. Current eye research. 2016;41(11):1492–1497.

12. Alten F, Clemens CR, Heiduschka P, Eter N. Localized reticular pseudodrusen and their topographic relation to choroidal watershed zones and changes in choroidal volumes. Invest Ophthalmol Vis Sci. 2013;54(5):3250–3257.

13. Rudolf M, Malek G, Messinger JD, Clark ME, Wang L, Curcio CA. Sub-retinal drusenoid deposits in human retina: organization and composition. Exp Eye Res. 2008;87(5):402–408.

14. Zweifel SA, Spaide RF, Curcio CA, Malek G, Imamura Y. Reticular pseudodrusen are subretinal drusenoid deposits. Ophthalmology. 2010;117(2):303-312.e301.

15. Curcio CA, Messinger JD, Sloan KR, McGwin G, Jr, Medeiros NE, Spaide RF. Subretinal drusenoid deposits in non-neovascular age-related macular degeneration: morphology, prevalence, topography, and biogenesis model. Retina. 2013;33(2):265–276.

16. Wang J, Jiang J, Zhang Y, Qian YW, Zhang JF, Wang ZL. Retinal and choroidal vascular changes in coronary heart disease: an optical coherence tomography angiography study. Biomed Opt Express. 2019;10(4):1532–1544.

17. Mano F, Sprehe N, Olsen TW. Association of Drusen Phenotype in Age-Related Macular Degeneration from Human Eye-Bank Eyes to Disease Stage and Cause of Death. Ophthalmol Retina. 2020.

18. Sawa M, Ueno C, Gomi F, Nishida K. Incidence and characteristics of neovascularization in fellow eyes of Japanese patients with unilateral retinal angiomatous proliferation. Retina. 2014;34(4):761–767.

19. Flamendorf J, Agron E, Wong WT, et al. Impairments in Dark Adaptation Are Associated with Age-Related Macular Degeneration Severity and Reticular Pseudodrusen. Ophthalmology. 2015;122(10):2053–2062.

20. Alten F, Heiduschka P, Clemens CR, Eter N. Exploring choriocapillaris under reticular pseudodrusen using OCT-Angiography. Graefes Arch Clin Exp Ophthalmol. 2016;254(11):2165–2173.

21. Yao H, Hou C, Liu W, Yi J, Su W, Hou Q. Associations of multiple serum biomarkers and the risk of cardiovascular disease in China. BMC Cardiovasc Disord. 2020;20(1):426.

22. Lambert NG, ElShelmani H, Singh MK, et al. Risk factors and biomarkers of age-related macular degeneration. Progress in retinal and eye research. 2016;54:64–102.

23. Smith RT, Merriam JE, Sohrab MA, et al. Complement factor H 402H variant and reticular macular disease. Archives of ophthalmology. 2011;129(8):1061–1066.

24. Alexander SL, Linde-Zwirble WT, Werther W, et al. Annual rates of arterial thromboembolic events in medicare neovascular age-related macular degeneration patients. Ophthalmology. 2007;114(12):2174–2178.

25. Duan Y, Mo J, Klein R, et al. Age-related macular degeneration is associated with incident myocardial infarction among elderly Americans. Ophthalmology. 2007;114(4):732–737.

26. Keilhauer CN, Fritsche LG, Guthoff R, Haubitz I, Weber BH. Age-related macular degeneration and coronary heart disease: evaluation of genetic and environmental associations. Eur J Med Genet. 2013;56(2):72–79.

27. Wang J, Xue Y, Thapa S, Wang L, Tang J, Ji K. Relation between Age-Related Macular Degeneration and Cardiovascular Events and Mortality: A Systematic Review and Meta-Analysis. Biomed Res Int. 2016;2016:8212063.

28. Colijn JM, den Hollander AI, Demirkan A, et al. Increased High-Density Lipoprotein Levels Associated with Age-Related Macular Degeneration: Evidence from the EYE-RISK and European Eye Epidemiology Consortia. Ophthalmology. 2019;126(3):393–406.

29. Monés J, Biarnés M. Geographic atrophy phenotype identification by cluster analysis. British Journal of Ophthalmology. 2017;102(3):388–392.

30. Wong CW, Wong TY, Cheung CM. Polypoidal Choroidal Vasculopathy in Asians. J Clin Med. 2015;4(5):782–821.

